# Simulating a community mental health service during the COVID-19 pandemic: effects of clinician–clinician encounters, clinician–patient–family encounters, symptom-triggered protective behaviour, and household clustering

**DOI:** 10.1101/2020.04.27.20081505

**Authors:** Rudolf N. Cardinal, Caroline E. Meiser-Stedman, David M. Christmas, Annabel C. Price, Chess Denman, Benjamin R. Underwood, Shanquan Chen, Soumya Banerjee, Simon R. White, Li Su, Tamsin J. Ford, Samuel R. Chamberlain, Catherine M. Walsh

**Affiliations:** Cambridgeshire & Peterborough NHS Foundation Trust, Fulbourn Hospital, Cambridge CB21 5EF; Department of Psychiatry, University of Cambridge, Sir William Hardy Building, Downing Site, Cambridge CB2 3EB

## Abstract

**Background:** Face-to-face healthcare, including psychiatric provision, must continue despite reduced interpersonal contact during the COVID-19 (SARS-CoV-2 coronavirus) pandemic. Community-based services might use domiciliary visits, consultations in healthcare settings, or remote consultations. Services might also alter direct contact between clinicians.

**Aims:** We examined the effects of appointment types and clinician–clinician encounters upon infection rates.

**Methods:** We modelled a COVID-19-like disease in a hypothetical community healthcare team, their patients, and patients’ household contacts (family). In one condition, clinicians met patients and briefly met family (e.g. home visit or collateral history). In another, patients attended alone (e.g. clinic visit), segregated from each other. In another, face-to-face contact was eliminated (e.g. videoconferencing). We also varied clinician–clinician contact; baseline and ongoing “external” infection rates; whether overt symptoms reduced transmission risk behaviourally (e.g. via personal protective equipment, PPE); and household clustering.

**Results:** Service organization had minimal effects on whole-population infection under our assumptions but materially affected clinician infection. Appointment type and inter-clinician contact had greater effects at low external infection rates and without a behavioural symptom response. Clustering magnified the effect of appointment type. We discuss infection control and other factors affecting appointment choice and team organization.

**Conclusions:** Distancing between clinicians can have significant effects on team infection. Loss of clinicians to infection likely has an adverse impact on care, not modelled here. Appointments must account for clinical necessity as well as infection control. Interventions to reduce transmission risk can synergize, arguing for maximal distancing and behavioural measures (e.g. PPE) consistent with safe care.

## INTRODUCTION

### Infection control strategies for community teams

Many countries have reduced interpersonal contact to control infection during the COVID-19 pandemic. The UK implemented interpersonal distancing on 16 March 2020 and “lockdown” on 23 March; its National Health Service (NHS) has reduced non-critical work and moved towards videoconferencing and telephone assessments where possible (1). However, some face-to-face consultations remain necessary, including for urgent mental or physical health needs. For community-based health services – including physical care teams, community mental health teams (CMHTs), mental health crisis teams, and other urgent care services – this raises the important question of how to structure patient contacts and clinical teams to minimize infection.

COVID-19 is a respiratory infection transmitted directly by airborne aerosols/droplets from an infectious person and indirectly via contaminated objects (fomites) (2). Asymptomatic people may be infectious (2,3). Airborne transmission is reduced by asymptomaticity (4), physical distance, time, and personal protective equipment (PPE) (2), and increased by symptoms such as coughing and aerosol-generating procedures (2). Fomite transmission is reduced by handwashing, cleaning/disinfection, and time, which cause viral inactivation (2,5).

Should community services bring patients to a central base (e.g. clinic) or visit patients at home? As of 8 April 2020, UK guidance advised PPE only for contact with patients having suspected or confirmed COVID-19, rather than for all patients (6). Primary care and outpatient settings should segregate COVID-19 and other patients in time or space, and allocate staff to one group or the other where possible (2). If we assume that clinic settings have appropriate infection control procedures in place, such as physical separation and cleaning, then a key difference between this and home visits is exposure of the clinician to the home environment. This additional exposure includes other household members and fomites. Does this pose a risk of increased transmission to the clinician and wider community? Is this influenced by different models of service provision?

Other questions relate to clinician–clinician encounters, and the impact of clinical activity on disease transmission to patients and families. To what extent do clinician–clinician and clinician–patient encounters affect spread? This may be important not only for the infection of clinicians, with implications for healthcare capacity, but because clinicians may be in contact with more people than most in a time of “social distancing” and thus might have the potential to infect a relatively large number of patients – one aspect of concern about “super-spreading” (7). Clinicians may also work with patients particularly vulnerable to COVID-19.

### Modelling approach

Epidemiologists have modelled COVID-19 spread across populations, and social measures to reduce spread given limited critical care capacity (4). However, we were unable to find assessments of the impact of community consultation strategy or clinician-to-clinician contacts on disease spread (via PubMed to 29 March 2020 or via enquiries on 24 March to the NHS Sustainability and Transformation Partnership for Cambridgeshire). We therefore simulated a population of clinicians, patients, and patients’ families via agent-based modelling (8), seeding the population with an infectious disease with the approximate characteristics of COVID-19. We examined spread under different conditions of interpersonal contacts, representing alternative ways of organizing community health services for necessary appointments. We report these simulations and make suggestions for infection control strategies applicable to such services.

We took a broad approach because there are insufficient high-quality data to infer reliable parameters for many aspects of a complex model. Uncertainties include: the infectivity profile over time; airborne transmission rates by contact type and time; the risks of fomite (surface) transmission in different contexts; many details of the network of patient and household community contacts (including community mixing in rural and urban areas, the use of public versus private transport, and the mix of patient residence between private homes and care facilities); the proportion of people in patients’ households who must continue to work; the consequences of antigen and antibody testing; the likelihood of serious morbidity or death following infection; etc. Therefore, we used a standard infectious disease model and applied simple “service-level” manipulations, including appointment type (varying the degree to which clinicians met patients and their family directly) and whether clinicians met each other. We tested the robustness of these effects by varying other inputs to summarize a wide range of unknown factors, such as the baseline and ongoing infection rates, and a form of inter-patient association or clustering. Furthermore, clinical care has competing objectives: to deliver optimal healthcare, to safeguard patients and staff from infection, to ensure the service is robust to staff shortages, to provide continuity of care, etc. An action that improves infection control may have other adverse effects. We did not model a mixed objective function explicitly. Instead, we focus on infection rates (including clinician infection rates) and discuss other potential effects of the service-level manipulations examined.

## METHODS

### Experiment 1

#### Fixed patient and disease-process parameters

We used a “susceptible–exposed–infectious–recovered” (SEIR) model (9), modified to distinguish the symptomatic and infectious periods (**Figure 1A**), modelling individuals stochastically (appropriate for small populations). Everyone began “susceptible”. People were randomly assigned to become symptomatic (if infected) with probability *p*=0.67, based on influenza (10) and close to the 69% (confidence interval 46%–92%) estimated for SARS-CoV-2 (11). Otherwise, infection would lead to asymptomatic infectiousness. People became infectious 4.6 days after infection (4). They remained infectious for 7 days, an uncertain value based on (12,13); we used a simple model without temporal variation in infectivity during that time. If symptoms developed, they began 5.1 days after infection (4,14,15) and lasted 7 days (12). Recovery precluded re-infection (also uncertain).

**Figure 1.**
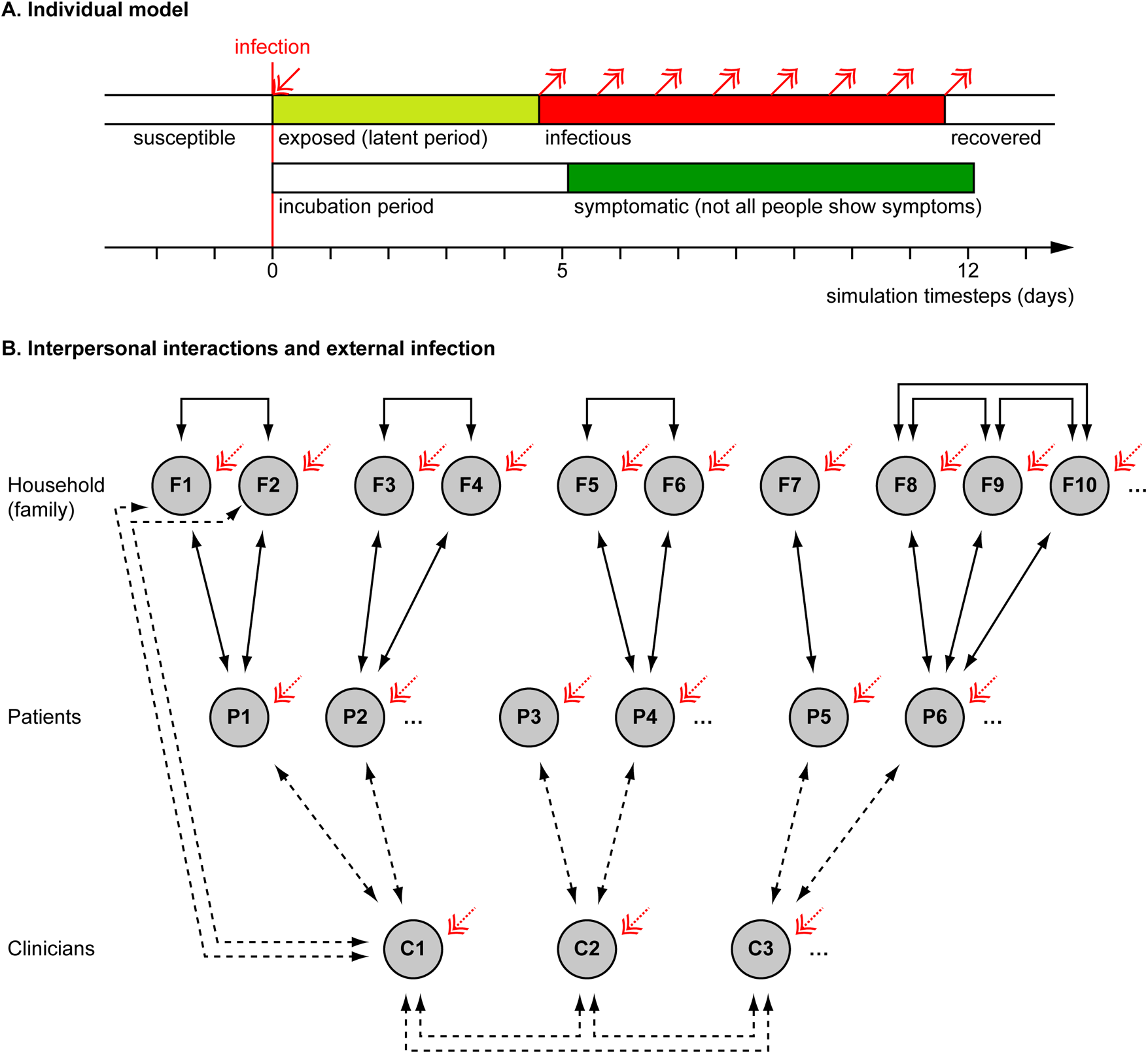
Overview of simulation methods. **(A)** Individual people followed a “susceptible–exposed–infectious–recovered” model, with full immunity after recovery. Whether an infected person developed symptoms or not was probabilistic (see Methods) and if they developed symptoms, this occurred slightly after the infectious period began. **(B)** Interaction between people (see Methods). Solid lines show fixed interactions; dashed lines show interactions that were varied. Patients interacted with their household (family) members and family members interacted with each other. Family sizes varied. In different conditions, clinicians did or did not interact directly with each other. Clinicians interacted with their patients, except in the “remote visit” condition. In the “family contact” (e.g. home visit) condition, clinicians also interacted briefly with family (not all such interactions drawn). The initial level of infection and the rate of ongoing “external” infection (red arrows) were varied. Additional manipulations, not shown here, included whether symptoms altered infectiousness in a behavioural, protective way (in contrast to an ever-present biological exacerbating way) and, in Experiment 2, whether patient households were grouped together in larger clusters with mutual interaction.

The transmission risk when a susceptible person was exposed to a symptomatic infectious individual was assumed to be *p*=0.2 for 24 hours’ exposure. We reduced this proportionally for shorter periods of exposure: a process with a constant rate of infection (or half-life *τ* for remaining uninfected), e.g. *p*_infection_(*t*) = 1 − 0.5*^t^*^/^*^τ^*, is approximately linear in this range. This value is uncertain: the basic reproduction number (*R_0_*) estimated for COVID-19 (16–18) implicitly incorporates the duration of infectiousness, the interpersonal contact rate, and the transmission risk per contact (itself depending on contact duration and transmission risk per unit time) (19). An *R*_0_ value of 2–3 (16) would be roughly equivalent to *p*=0.2 per 24h exposure if, for example, an infectious person infected 0.35 people/day for 7 days (2.5 people in total), e.g. via contact with susceptible others for 40 person-hours (e.g. 10 people, 4h each) per day. Asymptomatic people were 50% as infectious biologically as their symptomatic counterparts (4,10).

These values may be biologically inaccurate, and we did not implement realistic intersubject variability in these parameters. More sophisticated models exist (20). However, the absolute rate of infection was not our chief concern (as above); rather, we focused on the effect of service arrangements (described below). Relative differences due to service organization were assumed to be ordinally independent of the absolute transmission risk or other biological parameters.

#### Fixed population parameters

We simulated encounters between clinicians, patients, and patients’ household contacts (**Figure 1B**). A team of 20 clinicians was simulated, representing a multidisciplinary team (MDT). Each clinician saw 5 new patients per day, without follow-up appointments. Every clinician–patient interaction lasted 1h, based on a common UK “new patient” appointment slot in psychiatry. Patients were assumed to come from independent households. Patients could live with others (referred to here as family); the number of family members per patient was drawn from a Poisson distribution with λ=1.37, from the mean UK household size of 2.37 (21). Everyone in a household was assumed to interact daily and every household contact was assumed to last 8h/day. Clinicians’ households were not simulated, for simplicity (though the likely effect of adding clinician households would be to increase clinician infection rates slightly). People were assumed to follow household social isolation, i.e. with the exception of clinician encounters, there were no explicit household–household contacts. In addition, “external” infection was simulated (see below).

If a clinician became symptomatic, they were assumed to cease work for the duration of their symptoms and not meet others in the model during that time. Their appointments were reassigned to other clinicians (who therefore saw more patients, without limit). Symptomatic patients and family continued to interact with each other, and symptomatic patients were assumed still to require clinical care.

We simulated a population of mean size 14,240 (20 clinicians, 6,000 patients, and on average 8,220 family members) for 60 consecutive days (1–60, with no allowance for weekends). Days were simulated discretely; i.e. contacts were simulated (effectively) simultaneously for a given day, without regard to time of day.

#### Manipulations

The following aspects were varied, in all possible combinations. Variable ranges were chosen to represent plausible extremes.

- **Appointment type (“AT”)**. In the “patient only” (PO) condition, clinicians interacted only with the patient. This was intended to represent patients coming alone to a clinic, not interacting with others, with physical distancing and decontamination of clinic environments. In the “family contact” (FC) condition, clinicians also interacted for 0.2h (12 minutes) with each family member of the patient. This was intended to represent a home visit, but might also represent family accompanying the patient to a clinic and being present for a collateral history. In the “remote visit” (RV, e.g. videoconferencing) condition, no direct clinician–patient or clinician–family contact occurred.
- **Clinician–clinician meetings (“CM”)**. In the “clinician meeting” condition, all clinicians met up for 1h per day (e.g. for a handover or MDT meeting). In the “no clinician meeting” condition, clinicians did not interact with each other in person (e.g. used videoconferencing instead).
- **Baseline infection rate (“BL”)**. On day 0, the day before interpersonal contact simulation began, either 1% or 5% of the population were infected at random, reflecting approximate confidence interval (CI) extremes for the UK, 28 March 2020 (22). Clinicians and others had an equal chance of initial infection.
- **External infection (“EX”)**. Every person had a 0%, 0.5%, 1%, or 2% chance each day of becoming infected from external sources (e.g. in supermarkets, on public transport, etc.). The external infection rate is not constant in an epidemic, being affected by prevalence and the rate of external contacts; the upper figure of 2% was chosen to represent a very high value (e.g. if 10% of “external” people were infectious and each modelled person had 24 person-hours of “external” contact per day at transmission risk p=0.2 per person-day exposure as above).
- **Protective behavioural effect of symptoms upon transmission risk (“SX”)**. Being symptomatic has biological consequences (modelled above) but also social consequences. For example, overt illness may increase physical distancing or cause clinicians to use PPE, reducing infection risks. We chose values to represent plausible extremes of any such effect. In one condition, being symptomatic had an effect to reduce transmission risk to 10% of what it would otherwise have been (“behavioural symptom effect present”); the relative risk of infection with H7N7 avian influenza A is approximately 9% when using a respirator (23). Alternatively, the transmission risk was unmodified (“behavioural symptom effect absent”). If present, this effect applied to all interpersonal contacts (which may be unrealistic in that family members are unlikely to use PPE with each other; thus, any such behavioural effects may be smaller than those modelled).

#### Simulation

We simulated each condition 2000 times, using Python (https://www.python.org/).

#### Analysis

We used R (https://www.r-project.org/) to analyse the total number of (a) people and (b) clinicians infected by the end of the virtual experiment. We used a generalized linear model (GLM) with a Poisson distribution and a log link function, then analysis of deviance with type III sums of squares and *α*=0.05. However, the statistical power of a simulation is arbitrary (being determined by the effect size and the number of runs); thus, we present CIs and do not report exact *p* values, reporting instead “*p*<α” for 10^−3^≤*p*<0.05; “*p*≪*α*” for 10^−5^≤*p*<10^−3^; “*p*⋘*α*” for *p*<10^−5^. A significant interaction term implies non-additive effects of factors on the linear predictor, but because the predictors were on a log scale with respect to the dependent variable, interactions imply non-multiplicative effects of experimental factors on the number of people infected.

### Experiment 2

Not all patients live in small households: some live in much larger shared living facilities, such as care homes. Clustering might be used as a route out of “lockdown”. Does such patient clustering amplify any infection-transmitting effect of clinicians, who are unusually mobile people during times of “lockdown” and may travel between clusters? We examined whether the effects of appointment type and clinician meetings depended on the degree of patient clustering in such “hubs”, which we modelled as large households with multiple patients and their virtual “families” (or, similarly, care staff). We implemented this simply, by aggregating virtual households: thus, *n*=1 patients plus their family (household) per cluster as in Experiment 1, or *n*=2 households per cluster, etc. For simplicity, all co-residents were assumed to interact with each other, as patients and family would in a single-patient household, and clinicians were assumed to have “family-level” contact with everyone else in the cluster during FC-type visits. The selection of patients requiring a visit on any one day was random; those patients were then assigned cyclically to available (not sick) clinicians.

We varied the number of patients per household (“NPH”, 1−10, as a discrete predictor), AT, and CM. We held all other parameters constant at values suggested by Experiment 1 to give high power to detect such effects (no behavioural symptom effect; BL 1%; EX 0%).

### Deterministic SEIR model

To examine the basis of the synergy between interventions observed in Experiments 1–2, we ran deterministic plain SEIR models using the EpiDynamics R package. This form of model represents SEIR state changes via differential equations governed by rate parameters. It does not consider the timing, symptom, or interpersonal contact structure used in our agent-based model. We varied the proportion exposed at time *t=0* (1% or 5%, cf. Experiment 1) and the transmission rate *β*. Constants were: *t*_final_=1000 (for asymptote); birth/death rate μ=0; exposed-to-infectious rate *σ*=1/5; recovery rate *γ*=1/7. We examined the cumulative proportion infected (1 – susceptible).

## RESULTS

### Whole-population infection rates

In Experiment 1, whole-population infection was dominated by baseline and external infection rates (with infection spreading primarily via intra-household contacts), plus the behavioural response to symptoms (all *p*⋘*α*), with only very small contributions from the appointment type and clinician–clinician meetings (**Figure 2A**). That is, neither appointment type nor clinician meetings had any appreciable effect on the total number of people infected. Appointment type and clinician meetings had effects (e.g. AT×CM×BL×EX×SX interaction, *p*<*α*), but these effects were very small (the overall difference in the proportion infected was 0.01 percentage points between FC and RV conditions; **Figure 2A**). The beneficial effects of symptom-induced protective behaviour were proportionally greater in conditions with lower external infection rates (BL×EX×SX,*p⋘α;* **Figure 2A**).

**Figure 2.**
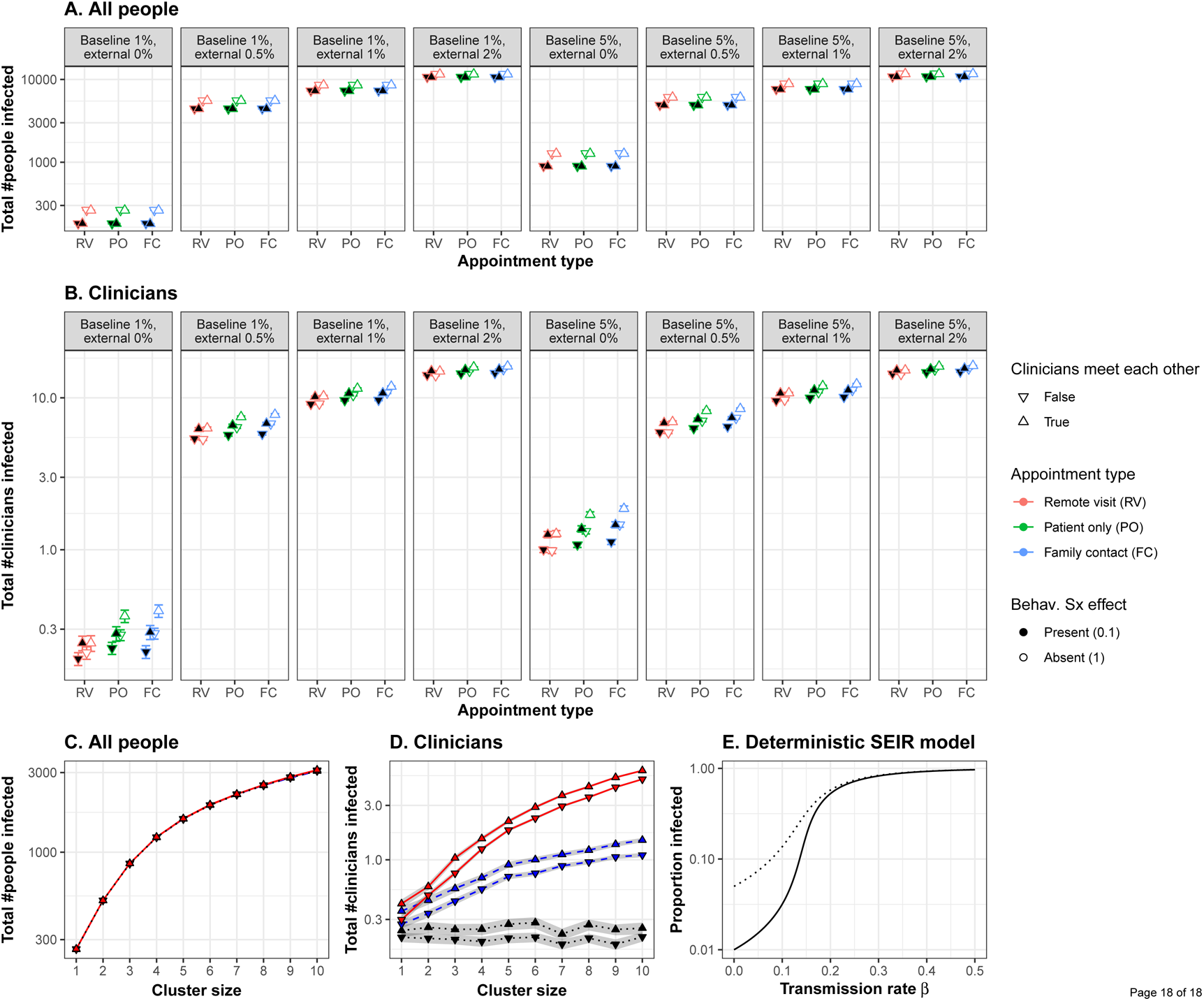
**(A)** Appointment type and clinician–clinician meetings had no substantial effect on whole-population infection. A protective behavioural response to symptoms had a proportionally greater effect at lower external infection rates. **(B)** Appointment type and clinician meetings had more substantial effects on the infection of clinicians. Moving to lower-contact appointment types and reducing clinician meetings showed synergy with a protective behavioural response to symptoms. **(C)** Clustering patient households together increased whole-population transmission. **(D)** Clustering increased clinician infection and substantially magnified the effect of appointment type. The beneficial effect of eliminating face-to-face clinician–clinician meetings was proportionally greater in low-clustering conditions, but this effect was very small. “Cluster size” refers to the number of patients (and their virtual families, or care home staff) per interacting “household”. **(E)** Changes in transmission rate have nonlinear effects on the total proportion of the population infected, shown in a deterministic standard SEIR model (solid line, 1% initially exposed; dotted line, 5% initially exposed). This effect underlies the synergy between measures that can be taken to reduce the transmission of infection: small changes in transmission risk sometimes cause dramatic changes in total infection. [RV, remote visits; PO, patient only; FC, family contact. Logarithmic scale. Error bars/ribbons are 95% CIs. “Behav. Sx effect” refers to the behavioural effect upon transmission when an infected person shows symptoms; either this reduces the transmission risk to 10% of its former value, e.g. via enhanced physical distancing or PPE, or has no effect.]

### Effects of appointment type on clinician infection

In contrast, appointment type had more substantial effects on clinician infection rates (**Figure 2B**), which depended upon external infection rates and the behavioural symptom effect (AT, *p*⋘*α;* AT×EX×SX, *p*⋘*α*). As expected from the number of contacts involved, infection rates were consistently ranked “family contact” (e.g. home visit) > “patient only” (e.g. clinic) > “remote” for all conditions. However, these effects were lessened at high external infection rates. We calculated the “new infection” rate as the final number of clinicians infected minus the number infected on day 0. We examined new infections in the PO condition as a proportion of the FC condition (PO new infections ÷ FC new infections) and similarly for RV versus FC. With 0% external infection, appointment type had a substantial effect (PO 82% of FC; RV 22% of FC); with 2% external infection, this was much less (PO 99% of FC, RV 95%). The effects of appointment type were larger without a behavioural symptom effect (**Figure 2B**).

### Effects of clinician–clinician meetings on clinician infection

There was a strong effect of clinician meetings, more pronounced at low levels of external infection (**Figure 2B;** CM, *p⋘α;* CM×EX, *p⋘α*). We calculated the effect’s magnitude as “new infections without clinician meetings ÷ new infections with clinician meetings” (“new infections” defined as before). The effect of eliminating clinician meetings was much larger with external infection at 0% (RV 0%, PO 36%, FC 38%) than at 2% (RV 94%, PO 95%, FC 95%), and numerically larger with a behavioural symptom effect (e.g. for 0% EX and PO appointments, eliminating clinician meetings cut new infections to 25% [of that with meetings] when there was a behavioural symptom effect, but only to 47% without that behavioural effect).

### Effects of symptom-related behaviour on clinician infection

Symptom-triggered behaviour had substantial effects on clinician infection rates (**Figure 2A,B;** SX, *p⋘α*; AT×EX×SX, *p⋘α*), as it did for whole-population rates. The beneficial effects of symptom-triggered behaviour were proportionally greater with lower external infection rates, for higher-risk appointment types, and without clinician meetings.

### Impact of patient clustering

**Figure 2C,D** shows Experiment 2’s results. Predictably, greater clustering increased infection rates (whole-population and clinicians, NPH, *p⋘α*). The effects of appointment type and clinician meetings on whole-population infection (AT×CM×NPH, *p⋘α*) were very small. The effects of appointment type on clinician infection were substantially magnified by greater clustering (AT×NPH, *p⋘α*).

### Synergy between service manipulations

Many of the effects of the modelled variables upon clinician infections were synergistic. Preventing physical clinician meetings had a greater proportional effect when external infection rates were lower, and with a behavioural protective response to symptoms (as above). Moving to lower-contact appointment types had a greater proportional effect when external infection rates were lower. Not all manipulations were synergistic (e.g. appointment types had a greater proportional effect without a behavioural response to symptoms, but both manipulations were nonetheless helpful). In a simple deterministic SEIR model, linear changes in transmission rate had nonlinear effects on the cumulative infection rate (**Figure 2E**), which is consistent with the synergies observed in the full model.

## DISCUSSION

### Summary

We modelled a hypothetical community clinical team, under different baseline and external infection rates. The fictional team organized its patient assessments in controlled environments in which only clinician–patient contacts occurred (e.g. managed clinics), visits in which some clinician–family contacts occurred (e.g. home visits or family present for a collateral history), or remote assessments in which the clinician and patient did not meet physically (e.g. videoconferencing). Clinicians met daily in person, or refrained from doing so. Under our assumptions, these service arrangements had only a very small impact on infection rates across the population studied (clinicians, patients, and family together; **Figure 2A,C**), but some had a substantial effect on clinician infection rates (**Figures 2B,D**). Clinicians may sometimes be a scarce resource, so higher clinician infection rates may have wider adverse effects on population health through lack of clinician availability. Behavioural measures to reduce transmission in response to overt symptoms also had substantial effects on infection rates, despite a period of “silent” infectiousness and some infectious people never exhibiting symptoms.

In our model, the infection risk to clinicians of appointment type directly reflected the degree of contacts with patients and family. This was expected, but we have quantified the relative importance of structural service manipulations. Eliminating daily face-to-face clinician–clinician meetings also had a noticeable effect on clinician infection rates, which was most pronounced, proportionally, with the lowest-risk patient encounter types. Combined risk reduction methods interacted with each other, sometimes having more than a multiplicative effect, and were disproportionately more effective than each alone. All these effects lessened with increasing rates of infection from outside the modelled population. Patient clustering increased whole-population infection and magnified the effect of appointment type on clinician infection, even for “patient-only” appointments. In our model, it was possible for multiple clinicians to visit a cluster; restricting which clinicians visit which clusters may be another important factor to consider. Clustering or “hub” effects may also occur in other ways not modelled here – such as patients being in receipt of multiple services from one or several provider organizations – and would similarly serve to increase transmission further. Changes in “lockdown” practices may affect external infection rates or clustering, requiring services to adapt to changing public health policy. Multiple small improvements in infection control can have a total effect greater than the sum of its parts (**Figure 2E**).

### Implications

These results emphasize and quantify an obvious point that minimizing contact with additional people, such as household contacts of a patient, contributes to infection control. Family contacts might occur during a home visit, but also if family accompany patients to a clinic. Videoconferencing or other remote assessment obviously provides the best infection control of the methods modelled here. However, there are trade-offs between infection control and clinical care for different appointment types, which must be judged by individual teams and clinicians. In particular, the choice between home visits and clinic assessments is complex and goes beyond the “family contact” aspects modelled here (**Table 1**). Other infection control differences include exposure to others during transport (likely favouring home visits by clinicians), and the risk of fomite transmission in either environment (hard to quantify, but potentially less predictable and greater for home visits). Relevant clinical differences go beyond infection control (**Table 1**).

**Table 1:** Infection control and clinical factors to consider for home visits, clinic appointments, and videoconferencing.

|  |  | Home visits | Clinic appointments | Videoconferencing |
| --- | --- | --- | --- | --- |
| Infection control | Positive | ▶ Patients remain at home and are not exposed to travel or to other patients or the public. | ▶ Able in principle to control environment, including distance between clinicians and patients, handwashing facilities, etc.<br>▶ Able to limit number: no relatives present (not always possible, as below).<br>▶ Very unwell and chaotic patients may not abide by “rules” of interpersonal space, reducing the infection control benefits.<br>▶ May be easier and safer to complete assessment with a single clinician in a single visit, reducing contacts. | ▶ Eliminates person-to-person contact for the attendees. |
|  | Negative | ▶ Possible increased number of household contacts with staff (family members etc.).<br>▶ Potential for limited handwashing facilities compared to clinics.<br>▶ Possible fomite transmission via contamination of environment (not known, not controllable). | ▶ Patients have to travel to clinic, possibly on public transport or via ambulance, or with an escort, increasing exposure.<br>▶ Patients are brought from the community to a clinical space, perhaps near to hospital inpatient sites, with risk of contact with other patients, staff members, etc.<br>▶ Some (e.g. single parents with multiple children) might have to bring one or more others.<br>▶ There is a particular need to consider the needs of high-risk/vulnerable patients including those who may be especially anxious about leaving home. | ▶ Without appropriate cleaning, the electronic device may act as a local fomite. |
| Quality of care and safety | Positive | ▶ Increased engagement with the most severely unwell patients.<br>▶ Sometimes the only option (e.g. patients who are not engaging, assessments under the Mental Health Act or equivalent legislation, etc.).<br>▶ Increased clinical understanding of patient based on their surroundings can be invaluable, particularly with the most unwell or vulnerable.<br>▶ Input from carers and family is enhanced. | ▶ Significantly less travel time for staff (compared to home visits) increases efficiency at a time of significant staffing concerns.<br>▶ A safe and calm environment that may aid assessment for some patients.<br>▶ Access for staff to supportive colleagues (but note the benefits of physical distancing). | ▶ May increase clinician efficiency compared to clinic appointments (e.g. reduced time between appointments; potential for automatic transcription via voice recognition).<br>▶ May be seen by some patients as a desirable balance of interpersonal interaction and infection control.<br>▶ Carer/family communication possible as for home visits.<br>▶ Colleague support as for clinic visits. |
|  | Negative | ▶ Less easy to ensure safety of staff.<br>▶ Home visits add travel time for clinicians, decreasing efficiency.<br>▶ Some homes are difficult to assess a patient in.<br>▶ If there are safeguarding concerns it is sometimes hard for a child or vulnerable adult to speak freely at home. | ▶ It may be distressing or even at times unsafe to ask very unwell patients to travel (perhaps very long distances).<br>▶ It represents a change in established patterns of working for some services.<br>▶ Risk of losing engagement with some high-risk patient groups. | ▶ May be inferior to assessment in person in terms of nuance, body language, situational awareness, and rapport.<br>▶ May be impossible for some patients, particularly those who are severely unwell.<br>▶ Relies on computing infrastructure (clinician’s, patient’s, intervening networks) which may fail.<br>▶ May be unfamiliar, reducing efficiency.<br>▶ Safeguarding concerns as for home visits. |

A striking result was the degree to which clinician–clinician meetings affected clinician infection rates, in some cases synergistically with other infection control measures. Current UK guidelines (2) include patient segregation, and segregation of primary care staff for COVID-19 and other patients, but do not currently recommend the segregation of all clinical staff, or all staff, from each other as far as is possible. Reducing contact between staff may be practical. Dividing a clinical team into subteams may provide partial benefits (**Table 2**). Fomite transmission is also a risk to clinical teams: precautions available to clinical teams against fomite transmission overlap with but have some distinct elements from those for airborne transmission (**Table 2**).

**Table 2.** Some behavioural infection control strategies for clinical teams. These are not exhaustive and are not intended to supplant local or national guidance.

|  | Reducing airborne transmission | Reducing fomite transmission |
| --- | --- | --- |
| All interpersonal contact | ► Standard national precautions (including physical distancing and appropriate PPE). | ► Standard national precautions (including handwashing, PPE, and surface cleaning). |
| Clinician–clinician interactions | ► Remote communication (e.g. phone, videoconferencing).<br>► For interpersonal contact, split into subteams and avoid mixing subteams. (Example: One team of 20 clinicians can have ${}_{20}C_2 = 190$ pairwise contacts. Four subteams of 5 can have $4 \times {}_5C_2 = 40$ contacts.)<br>► Minimize the number of staff “on site” at any one time (24). | ► Remote communication.<br>► Subteams and staff minimization as for airborne.<br>► Uniform/scrubs worn only at work, removed and laundered at day’s end.<br>► One-way flow of clinicians from “clean to dirty” (25) across a day where patients must be seen in person (e.g. confirmed negative → not suspected → suspected → confirmed COVID-19). Areas at clinicians’ base also segregated by “cleanliness”. Once progressed, clinicians have no contact with those in a “cleaner” category. Environmental cleaning regularly and at day’s end. |
| Clinician–patient interactions | ► Remote assessment (e.g. videoconferencing) where possible. | ► Remote assessment where possible.<br>► “Clean to dirty” progression as above. |

### Study limitations

The biological parameters we took for COVID-19 spread were estimates from the published literature and in some cases are subject to high uncertainty; likewise our estimates of initial and ongoing infection rates. Disease parameters were constant across subjects, other than symptomaticity given infection, which was stochastic. A paucity of contacts outside the household is highly atypical but corresponds to current UK policy, if not necessarily universal practice. The behavioural effect of symptoms upon transmission risk was modelled in the same way for household contacts as for clinician–patient contacts, which is unrealistic in that clinicians are more likely to have access to PPE and rules mandating its use. Clinicians’ households were not modelled and would tend to increase clinician infection rates (particularly if clinicians share a household). We also modelled a series of one-off patient assessments; many patients, of course, are seen repeatedly by their clinical teams, or see many different healthcare teams routinely. However, all these aspects were constant across conditions.

More important are limitations relating to differences between conditions. In the “family contact” (e.g. home visit) condition, we made assumptions about the duration of contact with family members, including that this plausibly encompassed the degree of surface as well as airborne transmission, and we assumed clinicians were not fomite vectors for direct transfer of virus between homes. In the “patient only” (e.g. clinic) condition, we assumed full segregation and cleaning between patients, with no inter-patient virus transfer. In the “no clinicians meeting” condition, we assumed a lack of fomite transmission between clinicians. Any of these may be unrealistic. Fomite transmission in either situation would likely increase infection rates and alter the impact of appointment type upon infection. We make our source code open for others to test different assumptions.

### Conclusions

Staff segregation, as well as appointment type and additional protective measures when meeting overtly symptomatic people, may have important effects on COVID-19 transmission in community clinical teams. Infection control manipulations can synergize with each other, suggesting that maximal implementation of such measures should be adopted to the degree possible. Appointment types must nevertheless meet the clinical need as well as infection control guidelines.

## Data Availability

Source code is available at https://github.com/RudolfCardinal/covid.

## DECLARATION OF INTEREST

RNC consults for Campden Instruments Ltd and receives royalties from Cambridge University Press, Cambridge Enterprise, and Routledge. BRU is clinical director of the Windsor Unit at Fulbourn Hospital, which delivers clinical trials in dementia/mild cognitive impairment for academic and commercial organisations without personal benefit, and is the clinical lead for dementia for the UK National Institute for Health Research (NIHR) Clinical Research Network in the East of England. SRC consults for Promentis and Ieso Digital Health.

## FUNDING

RNC’s, SC’s, and SB’s research is supported by the UK Medical Research Council (grant MC_PC_17213 to RNC). SRC is funded by a Wellcome Trust Clinical Fellowship (110049/Z/15/Z). This research was supported in part by the NIHR Cambridge Biomedical Research Centre; the views expressed are those of the authors and not necessarily those of the NHS, the NIHR, or the Department of Health and Social Care.

## ACKNOWLEDGEMENTS

We thank Mai Wong, Ruaidhri McCormack, Petra Vertes, Hisham Ziauddeen, and Carol Brayne for helpful discussion.

## AUTHOR CONTRIBUTIONS

CMW and CD posed the question. RNC performed the computational modelling and analysis. RNC and CMS drafted the manuscript. All authors contributed to, edited, and approved the final manuscript.

## DATA AVAILABILITY

Source code is available at https://github.com/RudolfCardinal/covid.

